# Impact of a Decision aid on Perceptions of Shared Decision-Making in the Primary Care Management of Patients with Subacromial Pain Syndrome: a two-phased multi-methods study

**DOI:** 10.1101/2025.02.14.25322309

**Authors:** Samantha C Bengtsen, Michael S Rathleff, Joshua R Zadro, Jens L Olesen, Nadine E Foster, Janus L Thomsen, Glyn Elwyn, Jens Søndergaard, Kristian D Lyng

**Author notes:** **Corresponding auth**or Kristian Damgaard Lyng, Department of Health Science and Technology, Aalborg University, Selma Lagerlöfs Vej 249, 9260 Gistrup, Denmark.

## Abstract

**Background:** Subacromial pain syndrome (SAPS) is the most common shoulder pain condition in primary care). Although exercise is the recommended first-line treatment, the absence of a universally superior option underscores the need for personalized care facilitated by shared decision-making (SDM). Despite the importance of SDM is increasingly recognized, its application in SAPS care remains poorly understood. The primary aim of this study was to explore the influence of a decision aid on patient and observer perceptions of SDM in the primary care management of patients with SAPS. Furthermore, the study aimed to explore correlations between patients’ and observers’ ratings of SDM.

**Methods:** We conducted a multi-methods study including observations of consenting patients with SAPS in their clinical consultations with clinicians from four Danish primary care practices, using OPTION-12. Additionally, we gathered patients’ perceptions of SDM two weeks after the consultation using an online survey using the 3-item CollaboRATE questionnaire and the 9-item Shared Decision-Making Questionnaire (SDM-Q-9). We observed consultations both with and without the introduction of a decision aid tailored to support the management of patients with SAPS.

**Results:** Thirty-four consultations were observed (16 with the decision aid, 18 without). Without the decision aid, the mean (SD) OPTION-12 score was low 10.5 + 3.3, and the median (IQR) CollaboRATE and mean (SD) SDM-Q-9 scores were 5 (IQR = 1.3) and 22.2 + 7.5, respectively. We observed higher scores in the consultations with the decision aid; the mean (SD, range) OPTION-12 score statistically significantly increased to 22.7 (6.87, 5-32), and the median (IQR) CollaboRATE and mean (SD) SDM-Q-9 scores were 6.5 (1.4) and 30.6 + 8.4, respectively. There was a positive and statistically significant correlation between patients’ and observers’ ratings of OPTION-12 and SDM-Q-9 scores across both phases. No significant correlation was found between CollaboRATE, OPTION-12, and SDM-Q-9 in either phase.

**Conclusion:** A decision aid significantly improved observer- and patient-rated SDM in primary care consultations for patients with SAPS. Observer-rated SDM scores more than doubled with the decision aid, and patients reported higher levels of SDM. These findings highlight the potential of decision aids to enhance SDM in SAPS care.

## Introduction

Subacromial pain syndrome (SAPS) is the most common shoulder pain condition in primary care [1]. Exercise is often used as first-line of care, but recent evidence has highlighted the lack of a definitive superior treatment strategy for managing SAPS [2–4]. Both in general, but especially in absence of superior treatments, it is important to involve patients in treatment decision-making, ensuring that their values and preferences guide the choice of treatment options [5,6]. Shared decision-making (SDM) is an approach to actively involve patients in decisions related to tests, treatments, and overall healthcare management [7]. Successful use of SDM includes a dialogue where the individual healthcare practitioner and patient use a collaborative approach to identify the most suitable treatment based on clinical presentation and patient preferences, while also considering the most relevant evidence [7,8].

Our recent needs assessment study highlighted that care-seeking patients with SAPS often require more knowledge and information to participate in SDM [9]. In this regard, decision aids are often considered as a sensible solution to accommodate such decisional needs [10]. Decision aids are clinical tools, often combining text and visuals, designed to present unbiased information on the pros and cons of different treatment options [11]. Recent evidence suggests decision aids have the potential to reduce decisional conflict, particularly feelings of being uninformed or uncertain about personal values [10]. However, limited knowledge exists on the practical use of decision aids in managing musculoskeletal pain and their impact on SDM.

We have developed a new decision aid which was initially evaluated in care-seeking patients with SAPS [12]. Although the study showed promising results in reducing decisional conflicts and regrets among patients, there remains limited knowledge about how SDM is currently utilised in clinical practice and whether decision aids can influence practitioner behavior. Therefore, the primary aim of this study was to explore the influence of a decision aid on patients’ and observers’ perceptions of SDM in the primary care management of patients with SAPS. Secondarily, we sought to explore correlations between patients’ and observers’ ratings of SDM, providing insights into the congruency of perceptions on SDM practices in primary care.

## Methods

### Design

We conducted a two-phased multi-methods study in primary care with patients with SAPS. The observations were conducted as a part of patients’ first consultation with general practitioners and physiotherapists. To explore changes in SDM, the observations were conducted in two distinct phases. Phase 1, referred to as the baseline observations, involved the observations of consultations without a decision aid (i.e., usual care). In phase 2, the intervention phase, observations were made during consultations where the decision aid was applied (**Figure 1**).

**Figure 1.**
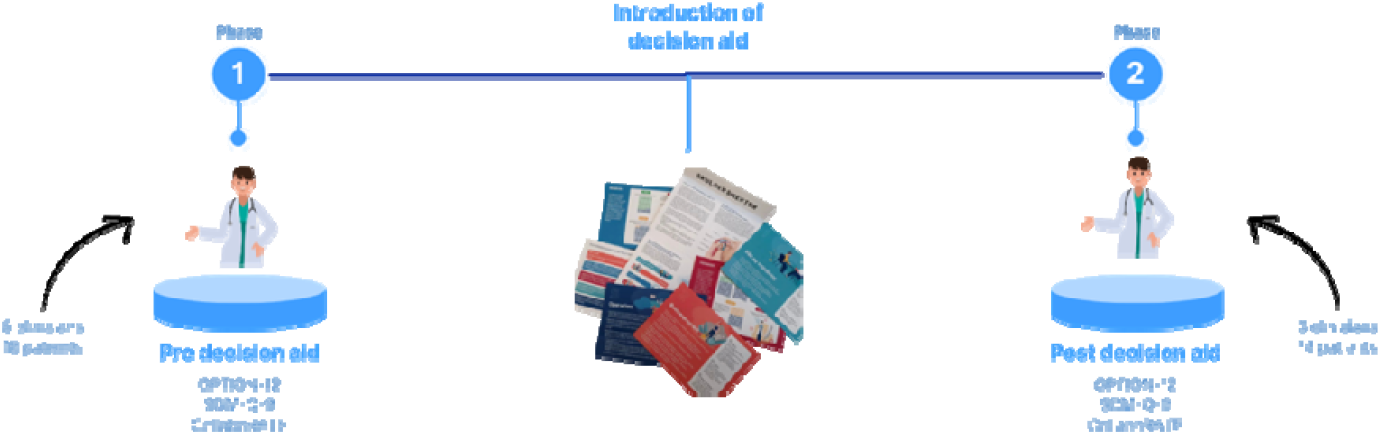
Study process. Observing Patient Involvement in Decision-making (OPTION)-12. Shared Decision-Making Questionnaire (SDM-Q9).

### Ethical considerations

Informed written and oral consent was obtained from all participants before the observations. The findings are presented in an anonymous manner, compliant with ethical standards to safeguard participant confidentiality. The study was conducted in accordance with the principles outlined in the Declaration of Helsinki and was considered exempt from full ethical review by the North Denmark Region Committee on Health Research Ethics (reference number: 2023-000206).

### Observer characteristics

SCB is a physiotherapist with a master’s degree in musculoskeletal physiotherapy, seven years of clinical experience managing patients with SAPS and experience undertaking qualitative research [9,12]. KDL is a physiotherapist and PhD candidate with experience in musculoskeletal pain research (20+ peer-reviewed articles) including qualitative research, however only with little knowledge on ethnographic studies (full description of the author group can be seen in **Appendix 1**).

### Context - previous development of a patient decision aid

We previously developed a decision aid containing information about invasive and non-invasive treatment options commonly used for patients with SAPS [12]. Further details on the development process, as well as alpha and beta testing, are thoroughly described elsewhere [12]. Prior to phase 2, All healthcare practitioners received a brief introduction to the decision aid involving information on its development (e.g., steps of development, end-user involvement, rationale for its development), the content (e.g., cards regarding how to use the decision aid, and option cards) and contact person (SCB or KDL).

**Figure 2.**
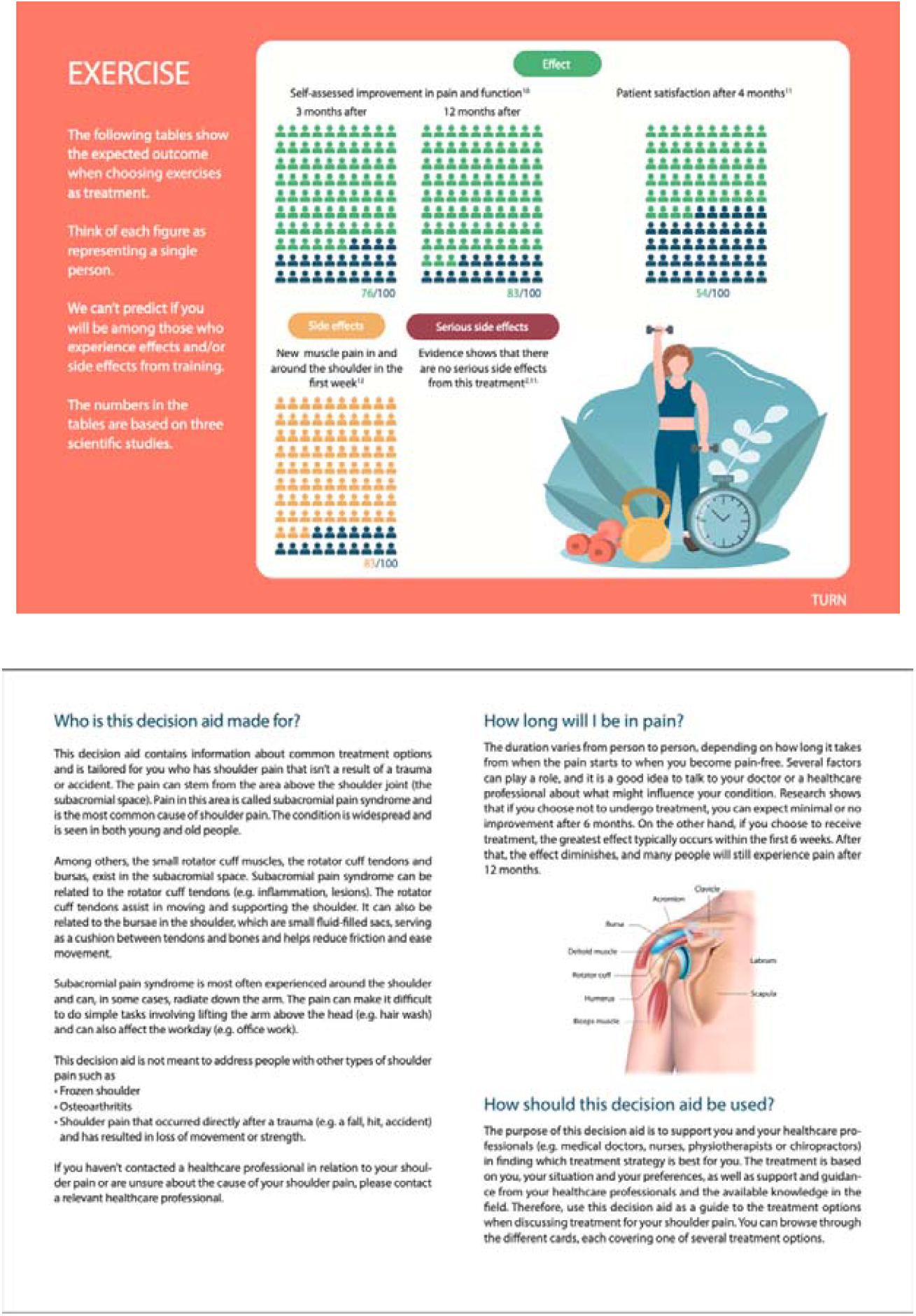
Example of decision aid for care-seeking patients with SAPS.

### Setting

Observations were conducted at two private physiotherapy clinics and two general practices in the North Denmark Region. Significantly differing in their daily practices, the physiotherapy clinics allocate 45-60 minutes per patient for the first consultation. In contrast, at the general practices, patient appointments are typically between 10-15 minutes. Patients at the physiotherapy clinics are typically referred by general practitioners, hospital specialists or healthcare insurers, although they can also self-refer to physiotherapy. General practitioners serve as the primary point of contact for patients experiencing non-traumatic shoulder pain, with financial coverage from the public healthcare system in Denmark (17). Unlike visits to general practitioners, patients in Denmark may have direct access to physiotherapists but typically incur out-of-pocket costs for physiotherapy services, including treatment or additional tests, unless these are covered by public health insurance or specific exemptions.

### Participants and sampling strategy

The target population for this study was care-seeking patients living with SAPS and healthcare practitioners involved in the management of these patients (e.g., general practitioners, physiotherapists). Healthcare practitioners were included if they possessed an authorized health license allowing them to manage patients with SAPS. Additionally, they had to have a minimum of two years of experience in addressing pain-related issues and manage at least five cases of SAPS annually (on average). Healthcare practitioners were invited to participate through local professional networks and emails. None of the healthcare practitioners were blinded to the study aim. Patients were eligible if they were 18 years of age or above and were suspected of having SAPS based on self-described symptoms or referral from general practitioners or healthcare insurers. Patients were included if they; were willing to participate and have their consultation observed, and if they could read and understand Danish. Healthcare practitioners were also required to provide consent prior to the observation of each consultation. Exclusion criteria encompassed traumatic onset of shoulder complaints or cases where SAPS could be definitively ruled out through clinical assessment of the patient’s healthcare practitioner. Furthermore, patients were excluded in the presence of symptoms indicating potential serious pathology (e.g., infection, malignancy, neurological conditions), and known severe psychiatric or psychological disorder.

### Data collection

The observations of patient-health care professional consultations at physiotherapy clinics and general practices in both phases of the study were conducted, in-person, by members of the research team (KDL and SCB) [13]. We collected data on gender, age, and educational level. To explore the influence of the decision aid on patient perceptions of SDM in the primary care management of patients with SAPS, we used the Observing Patient Involvement in Decision-making (OPTION-12) instrument [14]. OPTION-12 is a validated tool designed to measure the extent and quality of SDM [14]. The tool comprises 12 items [14,15]. Each item is rated on a 5-point scale from 0 to 4, with 4 indicating the highest level of competence and communication behavior in SDM. Consequently, the ratings for a consultation can range from 0 to 48 points. Both researchers thoroughly familiarized themselves with the OPTION manual beforehand, to ensure a consistent understanding of the scoring criteria. Subsequently, they jointly observed consultations with five patients, collaboratively scoring each consultation to ensure congruency and reliability in their assessments. Although a Danish version of the OPTION-12 instrument was not available at time of testing, both researchers were sufficiently proficient in English, allowing them to use the OPTION-12 instrument and manual in English comfortably. OPTION-12 has since been translated and culturally adapted to Danish [16]. To address the second aim of the study, we used SDM-Q-9 and CollaboRATE tools to measure patients’ perceived degree of SDM [17,18]. Patients were asked to fill out the questionnaires two-weeks after the consultation through a secure survey link using the web-application, REDCap^™^. A Danish version of both SDM-Q-9 and CollaboRATE tools were used, but only the SDM-Q-9 was culturally validated [19]. The SDM-Q-9 consists of nine items, rated on a Likert scale from 0 (strongly disagree) to 5 (strongly agree). CollaboRATE involves three questions, each item is scored from 0 (no effort was made) to 9 (every effort was made).

### Data analysis

We used descriptive statistics to determine means and standard deviations of demographics and outcomes for normally distributed data, and median and interquartile range (IQR) for non-normally distributed data. Normality of the data was assessed using visual inspection of Q-Q plots. Observer-rated SDM (OPTION-12) and patient-reported SDM (CollaboRATE and SDM-Q9) scores were compared between phases using appropriate statistical tests (independent t-tests or Mann-Whitney U tests) dependent on the normality of the data. Correlations between OPTION-12, CollaboRATE, and SDM-Q9 were assessed using correlation analysis (Pearson’s or Spearman’s correlation dependent on normality). Correlation strength was computed using an online Fisher’s r-to-z test calculator (http://vassarstats.net/rdiff.html). SPSS 25 for Windows (IBM Corp., Armonk, New York) was used for all analyses, with statistical significance set to p < 0.05 (two-sided).

## Results

### Participants characteristics

Sixteen local practices (eight physiotherapy practices, six general practices, two chiropractor practices) were invited, and four practices agreed to participate. From these, nine healthcare practitioners and 34 patients agreed to participate. Among the healthcare practitioners, seven were physiotherapists across two physiotherapy clinics, one was a physiotherapist in a general practice, and one was a general practitioner. Seven healthcare practitioners participated in phase 1, and six healthcare practitioners participated in phase 2 (of which four also participated in phase 1). Two patients declined to participate because of a lack of interest in participating, and four patients were excluded because of an ineligible diagnosis. In the first phase, 18 baseline observations were conducted at two physiotherapy clinics and one general practice. These encompassed 14 consultations conducted by six different physiotherapists and four consultations by one general practitioner. In the second phase, when the decision aid was included in the consultations, 16 observations were undertaken. Of these, 13 were conducted in physiotherapy consultations, while three were carried out in general practitioner consultations. Demographics of patient participants across the two phases are reported in **table 1**.

**Table 1.**
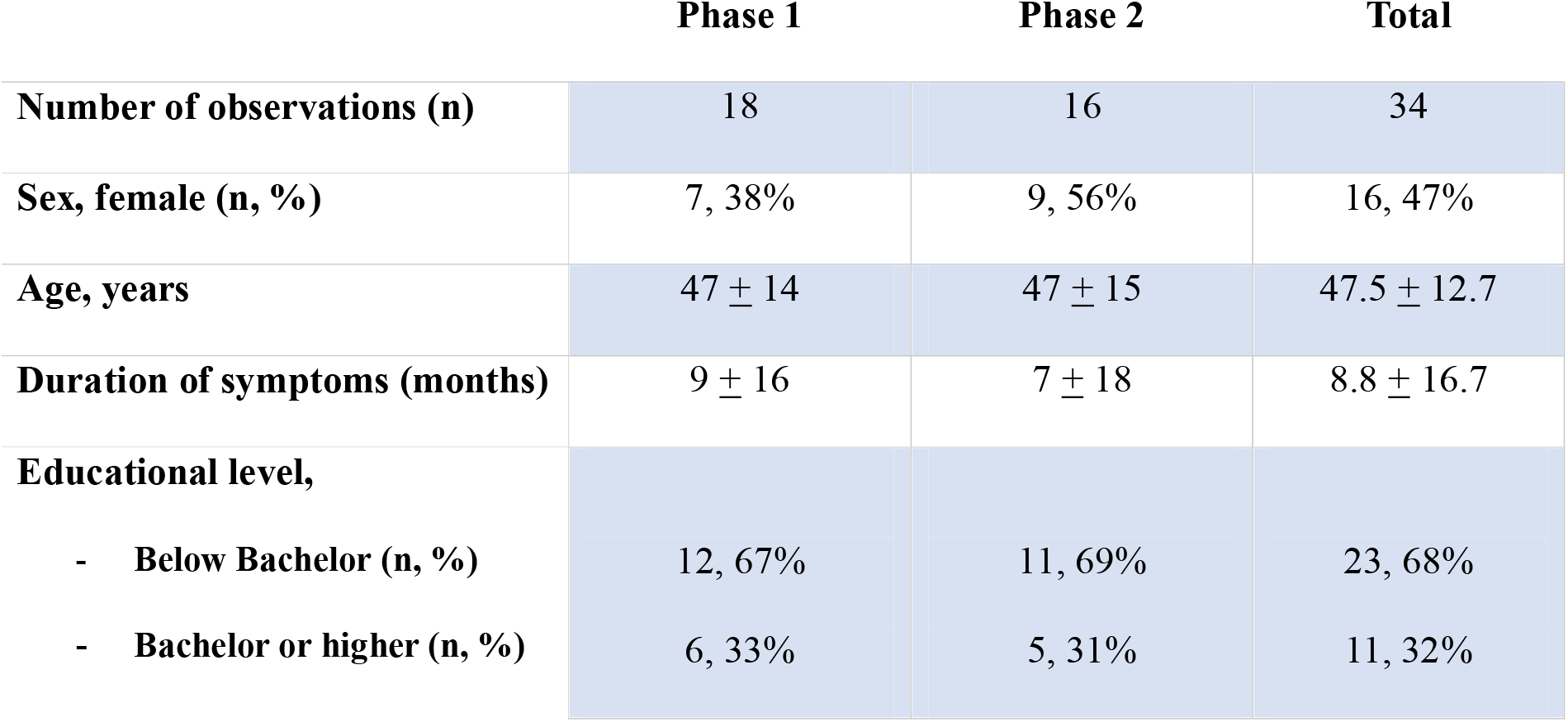
Patient Demographics across phase 1 and 2.

### Observers’ perception of shared decision-making

Visual inspection of Q-Q-plots indicated that data from the OPTION-12 total score did not violate the assumption of normality, thus parametric analyses were conducted. In phase 1, the mean (SD, range) total OPTION-12 score for all 18 consultations was 10.5 (3.3, 3-19) out of 48. In phase 2, a mean score of 22.7 (6.7, 5-32) was achieved. An independent sample t-test was conducted to compare differences in the OPTION-12 scores between the two phases. Assumption of equal variances between the two phases and outcomes was tested using Levene’s test, and the results showed no significant differences in variance (OPTION-12, *F*= 2.967, *P* = 0.095). Results indicated a statistically significant difference between the two phases in OPTION-12 scores, t(32) = 6.8, *P* = <0.001. The mean difference between the phases in OPTION-12 scores was 12.2 (95% CI [8.6, 15.8]), indicating significantly higher SDM scores in consultations in phase 2. See **Table 2** for mean scores and change-scores between the two phases.

**Table 2.** Change scores of OPTION-12, SDM-Q-9, and CollaboRATE across phases. *Statistically significant † Non-normally distributed, thus statistics is reported using median, IQR, and Mann-Whitney U statistics.

|  | Phase 1 | Phase 2 | Difference<br>score | Test-type | P-value | 95% CI |
| --- | --- | --- | --- | --- | --- | --- |
| **OPTION-12** | 10.5 ± 3.3 | 22.7 ± 6.7 | 12.2 ± 1.7 | T-test | <0.001* | [8.6, 15.8] |
| **SDM-Q-9** | 22.2 ± 7.5 | 30.6 ± 8.4 | 8.4 ± 2.7 | T-test | 0.004* | [2.8, 14.1] |
| **CollaboRATE**<br>† | 5, IQR = 1.3 | 6.5, IQR = 1.4 | 30.0 | Mann-Whitney U | <0.001* | - |

### Patients’ perception of shared decision-making

Normality testing using the Q-Q-plots test indicated that data from the SDM-Q-9 total score did not violate the assumption of normality, while data from the CollaboRATE total score deviated from normality. Given these findings, an independent samples t-test was conducted to compare SDM-Q-9 scores between the two phases, while a Mann-Whitney U test was applied to compare CollaboRATE scores. In phase 1, the mean total SDM-Q-9 score for all 18 consultations was 22.2 (7.5, 1-37) out of 45. The median score across items for CollaboRATE was 5 (IQR = 1.3) out of 10. In phase 2, the mean total SDM-Q-9 score for all 16 consultations was 30.6 (8.4, 5-40). The median CollaboRATE score was 6.5 (IQR = 1.4). The results from Levene’s test showed no significant differences in variance for SDM-Q-9 scores (*F*= 0.063, *P* = 0.803). The independent samples t-test indicated a statistically significant difference between the two phases in SDM-Q-9 scores; t(32) = 3.0, *P* = 0.004, with a mean difference of 8.4 points (95% CI: [2.8, 14.1]. Results from the Mann-Whitney U-test showed a significant difference between the two phases, U = 30.0, Exact *p* = <0.001. Together, these findings suggest that the decision aid increased SDM, as reflected by both the SDM-Q-9 and CollaboRATE measures. See **Table 2** for mean scores and differences between the two phases.

### Correlation between observed and perceived SDM

A significant positive correlation was found between OPTION-12 and SDM-Q-9 scores in the first phase (r = 0.865, *P* = <0.001), which remained significant in the second phase (r = 0.887, *P* = <0. 001). The Fisher’s r-to-z test revealed a statistically significant change in correlation strength between OPTION-12 and SDM-Q-9 between the two phases (Z = 3.23, *P* = 0.001). This finding suggests that the relationship between these two measures of SDM was improved in phase 2. A Spearman correlation revealed a non-significant correlation between CollaboRATE and OPTION-12 scores in phase one (ρ = 0.340, *P* = 0.168). This correlation remained non-significant in phase two (ρ = 0.218, *P* = 0.417). Similarly, the correlation analysis showed a non-significant relationship between CollaboRATE and SDM-Q-9 scores in phase one (ρ = 0.314, *P* = 0.204). The correlation was also non-significant in phase two (ρ = 0.110, *P* = 0.686). See **Table 3** for correlation between OPTION-12, SDM-Q-9, and CollaboRATE across phases.

**Table 3.** Correlation between OPTION-12, SDM-Q-9, and CollaboRATE across phases. *Statistically significant

| *Variable(s)* | *Phase* | *Correlation coefficient* | *P-value* | *N* |
| --- | --- | --- | --- | --- |
| OPTION-12 & SDM-Q-9 | Phase 1 | $r = 0.865$ | $<0.001^*$ | 18 |
| | Phase 2 | $r = 0.887$ | $<0.001^*$ | 16 |
| OPTION-12 & CollaboRATE | Phase 1 | $\rho = 0.340$ | 0.168 | 18 |
| | Phase 2 | $\rho = 0.218$ | 0.417 | 16 |
| SDM-Q-9 & CollaboRATE | Phase 1 | $\rho = 0.314$ | 0.204 | 18 |
| | Phase 2 | $\rho = 0.110$ | 0.686 | 16 |

## Discussion

### Summary of findings

This study explored the influence of a decision aid on patients’ and observers’ perceptions of SDM in the primary care management of patients with SAPS. Also, we explored correlations between scores on SDM from consultations without (phase 1) and consultations with the use of our decision aid (phase 2). The study highlighted higher SDM scores in phase 2, where the decision aid was used as a tool for the consultations which were present from both observer’ and patient’ ratings. A significant positive correlation was found between OPTION-12 and SDM-Q-9 scores, with the strength of this relationship increasing in phase 2. This suggests greater alignment between observed healthcare practitioner behaviors and patients’ perceptions of SDM in phase 2. No significant correlations were found between CollaboRATE scores and either OPTION-12 or SDM-Q-9 scores across phases. This surprising finding may indicate that CollaboRATE captures different aspects of SDM that are less directly tied to those measured by OPTION-12 and SDM-Q-9. This discrepancy highlights the complexity of assessing SDM and the potential need for multimodal measurement approaches.

### Comparison with previous research

The available evidence on how decision aids impact observed shared decision-making behaviors in pain conditions is limited [10,20,21]. Consistent with our findings, Bowen et al. (2020) reported that decision aids improve patients’ understanding of treatment options for chronic musculoskeletal pain [20]. However, their review highlighted that while decision aids enhance knowledge, their impact on broader decision-making processes and patient satisfaction is variable. This may suggests that additional factors, such as healthcare practitioner communication, may influence their overall effectiveness. Our study suggests that SDM can be influenced by a decision aid, however more research is needed. Our decision aid was partly adapted from a recent online randomized controlled trial which found that a decision aid targeting people with SAPS on a waitlist considering shoulder surgery, had no effect on treatment intention, attitudes, informed choice, and decisional conflict [22]. The lack of effect where mainly attributed towards timing and delivery-method of the decision aid [22]. Our adapted version, which were delivered in the earlier decisional stages and by a healthcare practitioner in primary care, showed more promising results in terms of influence on decisional conflicts [12]. These findings shed light on the possible importance of understanding the contextual- and patient-specific characteristics that may influence the effectiveness of decision aids and SDM perceptions. Our findings suggest SDM-Q-9 scores were higher in consultations using a decision aid. However, the magnitude of improvement in SDM-Q-9 scores observed in our study exceeds what has been reported in most studies, where little-to-no differences have been reported between randomized controlled trials were interventional groups had received training in SDM compared to no training [23]. This discrepancy could be attributed to differences in the study design, patient characteristics (e.g., level of decision-preparedness), or the complexity of decisions addressed. Furthermore, our results might be partly explained by the fact that our decision aid was developed for both patients and healthcare practitioners. The evidence for effectiveness of multi-faceted interventions is, however, inconclusive, and further evidence is needed to understand the impact of such strategies [24,25]. Our study also showed significant improvements in CollaboRATE scores after the introduction of a decision aid, however the scores were not correlated with SDM-Q-9 or OPTION-12. Unlike the OPTION-12 and SDM-Q-9, which focus on specific clinician behaviors and structured decision-making processes, CollaboRATE reflects patients’ subjective experience of involvement. These distinct differences may explain the lack of correlation and further underscore that CollaboRATE likely assess a unique dimension of SDM, offering a complementary perspective when evaluating SDM behaviors. Similarly to our study, Ubbink et al. conducted a secondary analysis combining data from five studies (n = 422 patients) across various outpatient and inpatient departments, comparing CollaboRATE and SDM-Q-9 scores [26]. They found a moderate correlation between the two measures across diverse patient groups, including those from congenital vascular malformations, anesthesiology, cardiology, nephrology, radiotherapy, psychiatry, internal medicine, and hematologic oncology, spanning four different university medical centers or hospitals in the Netherlands [26]. The study also found a clear ceiling effect across both measures, but the effect was much stronger for the CollaboRATE tool [26].

Together with our findings, these results highlight the need for future studies to investigate which measures work for whom, when and why, and the importance of including both subjective and objective measures of SDM.

### Implications for future research

We co-developed a decision aid with key stakeholders to support SAPS patients in primary care [9,12]. While our findings suggest a positive impact on SDM, a randomized controlled trial is needed to compare its use with usual care in a larger and more diverse population. The discrepancy between CollaboRATE and other SDM measures highlights a key research gap. Additionally, the long-term effects of decision aids in musculoskeletal care remain unclear. Their effectiveness depends on clinician SDM competency, yet many lack formal training [27,28]. In our study, minimal training may have influenced results. Future research should assess the impact of structured SDM training and whether additional strategies are needed to sustain improvements. Finally, studies should examine how SDM evolves over time and its effects on clinical outcomes like pain, function, and quality of life.

### Strengths and limitations

The use of observational compared to patient-reported methods assessing SDM as it occurs in clinical practice increase the applicability and relevance of our findings to clinical practice in primary care. Furthermore, the study is strengthened by the use of multiple validated tools to assess SDM. This multi-modal approach strengthens the robustness of the findings, as it captures complementary perspectives on SDM, offering valuable, although explorative insights into how decision aids impact both observers’ and patients’ ratings of SDM. There are several limitations of our study. The main limitations of the study are a small sample size and lack of randomization which should raise caution of the conclusion of this study. Secondly, observations were conducted by two observers who were not blinded to the hypothesis, and furthermore, didn’t audio or video record the observations. These limitations hamper both the consistency of the measurements (inter- and intra-reliability), and might also impose observer bias, which possibly over-inflate OPTION-12 scores [29]. Our study primarily recruited physiotherapists, and recruitment of healthcare practitioners was primarily based on authors’ personal networks. The lack of other healthcare practitioners will influence the generalizability of our findings, and the recruitment through personal networks might impose selection bias where most participants already had an interest in SDM. This study lacked data collection of healthcare practitioners’ perceptions of SDM due to practicality-issues in clinical practices. These data would have provided valuable additional insights. Lastly, in our study we collected data on patients’ perception of SDM two weeks after a consultation. This was done to minimize the burden on patients but might impose recall bias [30].

## Conclusion

Our study suggests that a decision aid can enhance both patients’ and observers’ ratings of SDM in primary care for SAPS. The positive correlation between these ratings indicates alignment between observed SDM and patient experience. However, the CollaboRATE measure did not follow this pattern, suggesting it captures different aspects of SDM. Future research should validate these findings in larger trials and further explore differences between SDM measures.

## Data Availability

All data produced in the present study are not available.

