## Appendix 1 for "Impact of a Decision aid on Perceptions of Shared Decision-Making in the Primary Care Management of Patients with Subacromial Pain Syndrome: a two-phased multi-methods study"

**Appendix 1. Researcher characteristics**

MSR, JRZ, JLO, NEF, JLT, GE, and JS supervised the research project. MSR is a professor specializing in musculoskeletal health and implementation, with extensive experience in health research. JRZ is a physiotherapist and Senior Research Fellow with expertise in developing and evaluating decision aids. JLO is a professor and experienced rheumatologist with over 20 years of clinical experience managing SAPS. NEF is a professor, a physiotherapist and NHMRC Leadership Fellow with a focus on musculoskeletal pain, orthopaedic and primary care research including mixed methods studies. GE is a professor and a world-leading expert in shared decision-making and decision aids. Both JLT and JS are professors in family medicine and experienced general practitioners with significant contributions to primary care research.
